# The Application of Data Science Techniques and Algorithms in Women’s Health Studies

**DOI:** 10.1101/2022.03.27.22273006

**Authors:** Ikpe Justice Akpan, Obianuju Genevieve Aguolu

## Abstract

**Objective:** Evaluate and map data science methods employed to solve health conditions of women, examine the problems tackled and the effectiveness.

**Research Method:** Text analytics, science mapping, and descriptive evaluation of data science methods utilized in women-related health research.

**Findings:** (i). The trends in scholarships using data science methods indicate gaps between women and men relating to health burden and access to health. (ii). The coronavirus (SARS-CoV-2) outbreak and the ongoing COVID-19 pandemic tend to widen the identified health gaps, increasing the disease burden for women, while reducing access to health. There are noticeable additional health burdens on pregnant women and those with several health conditions (breast cancer, gynecologic oncology, cardiovascular disease, and more). (iii). Over 95% of studies using data science methods (artificial intelligence, machine learning, novel algorithms, predictive, big data, visual analytics, clinical decision support systems, or a combination of the methods) indicate significant effectiveness. (iv). Mapping of the scientific literature to authors, sources, and countries show an upward trend; 997 (16%), 113 (1.33%), and 57 (2.63%) per article, respectively. About 95% of research utilizing data science methods in women’s health studies occurred within the last four (4) years.

**Conclusions:** The application of data science methods in tackling different health problems of women is effective and growing, and capable of easing the burden of health in women. The ongoing COVID-19 pandemic tends to compound the health burden for women more than men. Policymakers must do more to improve access to health for women.

## 1. INTRODUCTION

The use of information systems, computer science methods, and operations research techniques to improve operations performance in the healthcare sector has a long history.^1-5^ Data science (DS) techniques and analytical methods are also employed to mitigate health disparities across social, cultural, and sex classification more recently.^6,7^ The goal is to eliminate the known gaps in healthcare access, reduce disease burden, and improve clinical outcomes and healthcare for all.^8,9^

The health gaps between men and women in terms of disease burden and access to health are well established in the social science and healthcare literature, including the ongoing coronavirus disease (COVID-19).^122^ Gaps are also reported during the ongoing coronavirus disease (COVID-19) pandemic regarding disease burden and access to health. While the pandemic places an additional health burden on women than men, such as pregnant women and those with pre-existing breast cancer, gynecologic oncology, and other health conditions, fewer women (on average) have access to health compared to men. For example, in studies reported in this paper, pregnant women diagnosed with COVID-19 can have severe morbidities and have greater mortality risk. Simultaneously, child-care responsibility and economic burdens rests on working mothers during lockdowns and working from home rests more.^45,74,100,103,122^

The recent advances in computer processing power and the availability of massive and complex data comes the use of DS tools and methods and big data analytics techniques to conduct healthcare and biomedical sciences research and practice.^10,11^ The ongoing advances in DS and analytics methods include machine learning, deep learning, artificial intelligence, which in addition to visual analytics and information visualization, enhances knowledge discovery, sensemaking, understanding, and offering solutions to complex medical and health problems.^11-15,108^ Recent studies identify healthcare research and practice as one of the active fields utilizing data science technologies and methods to enhance clinical decision support systems and improving health outcomes.^5,10,11,60^ Therefore, this study undertakes an evaluation of the data science techniques and novel algorithms employed in women’s health studies. We also map specific approaches to the type of problems solved, identify what works and what does not, and identify areas that require more work.

## 2. THEORETICAL BACKGROUND

This section examines the peculiarity of women’s health and the disparity in terms of the burden and access to health.

### 2.1 Women’s Health

Certain diseases and conditions primarily affect women due to differences in their body anatomy and physiology. Some peculiar health conditions that impact women include breast health, obstetric and gynecological issues. Others are and biocompatibility issues related to implants used in women.^17^ Furthermore, several factors can influence the way women respond to common diseases and medical products. These factors include intrinsic (age, genetics, hormones, body size, sex-specific physiology, pregnancy), extrinsic factors (diet, sociocultural issues, environment).

Sex-specific biomarkers vary in females by age because of variations in hormone level, co-morbidities, and taking multiple medications. The toxicity of some medications can increase due to changes in a woman’s physiology during pregnancy. Also, certain medications can be contraindicated in women during lactation due to potential toxicity or teratogenicity to the growing fetus, limiting their therapy choices. Some common diseases that may affect women differently include osteoporosis, autoimmune diseases, certain cancers such as breast, uterine, cervical cancers, cardiovascular diseases, lung diseases, neurological diseases, and certain psychiatric conditions.

Osteoporosis is one of the common health problems experienced by older women due to their physiology. Women experience fluctuations in estrogen throughout life. Depletion of estrogen level causes osteoporosis, a risk factor for fractures, increased morbidity, and mortality. Moreover, this condition negatively impacts postmenopausal women’s physical, emotional, and mental well-being.^18^ Thus, it is essential to consider women’s risk factors when providing care or managing cases.

The disparity in Women’s healt is also prevalent in autoimmune diseases. Those conditions include autoimmune thyroid, multiple sclerosis, and rheumatological systemic autoimmune. Women also carry more disease burden on systemic lupus erythematosus, Sjögren’s syndrome effect, and rheumatoid arthritis, and represent most of these cases.^8,19,45^ The reasons for this imbalance are unclear but attributed to genetic (X-linked) and hormonal factors. Furthermore, the women’s health imbalance is present in the course and prognosis of the diseases. More studies are needed to investigate if this imbalance is due to differences in the disease’s biology, how men and women respond to therapy or the treatment.^8,9,19,45^

Estrogen has anti-inflammatory effects. The fall in estrogen levels as women age predisposes them to arthritis. Older women are more likely to get arthritis than men of the same age. They are also more likely to experience worse pain from it more than men. They are also more vulnerable to rheumatoid arthritis, a highly debilitating form of arthritis. Furthermore, arthritis tends to affect women’s limb joints more than men due to pregnancy changes and female anatomy.

In the United States, cardiovascular diseases are the leading cause of death among women. Most of these deaths are caused by coronary heart diseases and are usually sudden. Coronary heart diseases are responsible for 20% of female deaths. About 1 in 16 women aged 20 and older (6.2%) have coronary heart disease, the most common heart disease.^20^ Framingham’s study, pioneering research in the sex differences, demonstrated that women have greater primary risk factors for heart diseases than men.^21^ Furthermore, studies found that women are more likely to have worse outcomes from heart diseases. Possible factors related to worse effects in women include delays in seeking medical care or voluntary discharge. The reasons for these delays are unknown. More studies should be directed at understanding these risk factors and should be made available to the public, particularly women.

There are sex-specific disparities in the public health burden of cancers due to variations at the genetic/molecular level and sex hormones such as estrogen.^22^ The genetic and hormonal differences influence gene expressions of certain cancers, drug metabolism, and therefore the effect of chemotherapy. These differences should be considered in cancer research to avoid disparity of chemotherapy’s efficacy and toxicity between sexes. Women are more likely to experience psychiatric disorders, alcohol abuse, substance use disorders, conduct disorder, syndromal adulthood antisocial behavior without conduct disorder,^23^ tranquilizer abuse with social and specific phobias,^24^ and major depressive disorder and anxiety disorders compared to men.^26^ Females are more likely to experience the double stigma of being both alcoholic and morally degenerate. They are more likely to face societal stigmas at work, home, school, community. Furthermore, women with mental health disorders and drug addictions experience more stigma than men.^25^ The biological difference is attributed to the sex chromosomes. Awareness and understanding of the sexual dimorphism in neuropsychiatric disorders, increased risk of neurodevelopmental and psychiatric disorders in sex-linked genetic disorders can guide the management of these disorders.^26^

### 2.2 Disparities in Women’s Access to Healthcare

Previous studies have identified sex disparities in access to care.^7,8,27^ While health needs are significantly greater among older women than men of the same age, women have fewer economic resources.^27^ Even after controlling for health needs, there are still sex differences in preventive care and increased differences in hospital services. Women are less likely to have hospital stays or outpatient services^28^ than men with similar demographic and health profiles. Women are more likely than men to report insufficient money for medications, delays in care,^29^ insufficient money for health care, and insufficient money for mental health care. Studies show that while women often develop drug and alcohol addictions faster than men, they are likely to face multiple barriers to substance use treatment and are less likely to see treatment.^25^ They are more likely to have poorer treatment outcomes because of more inadequate resources. Some factors that may contribute to a delay in women seeking psychiatric care include hesitancy to discuss reasons they started using substances, difficulties or fears associated with leaving familial roles, stigma. They are five times more likely to have a history of sexual abuse.

## 3. MATERIALS AND METHOD

### 3.1 Research Objectives

This study seeks to achieve the following four objectives:

RO1. Identify and analyze the temporal trends of DS applications in women’s health research.

RO2. Map specific DS techniques/algorithms to women’s health conditions addressed, evaluate the effectiveness, identify what works or does not, and determine alternative methods.

RO3. Examine the relationship between the scientific literature (SL) and usage and the citation impacts.

### 3.2 Data Collection

The data used in this study came from published documents selected through a literature survey performed on the scientific database index of the “ISI Web of Science (WoS) core collection.” The search covers the period 2000 and 2020, which double as the effective trending era of DS and big data analytics.^11^ Our initial literature survey for this study occurred on February 22, 2021, with the final update on March 3, 2021. The complete search terms are as shown in Table 1. Limiting the search and selection of representative sample documents to the WoS was to ensure the quality of the publications included in this study and weeds out papers from unreliable sources.^30^ This is particularly important for a trending topic like DS/big data and visual analytics that attracts a diverse research interest.^11,31^ However, all the indexed articles on the WoS core collection addressing the topic of interest had an equal chance of being selected according to the preferred reporting items for systematic reviews and meta-analyses: The PRISMA statement.^32^ Also, articles indexed on the WoS make up about 84% of the scholarship indexed in MEDLINE and SCOPUS.^31,33^

**Table 1.**
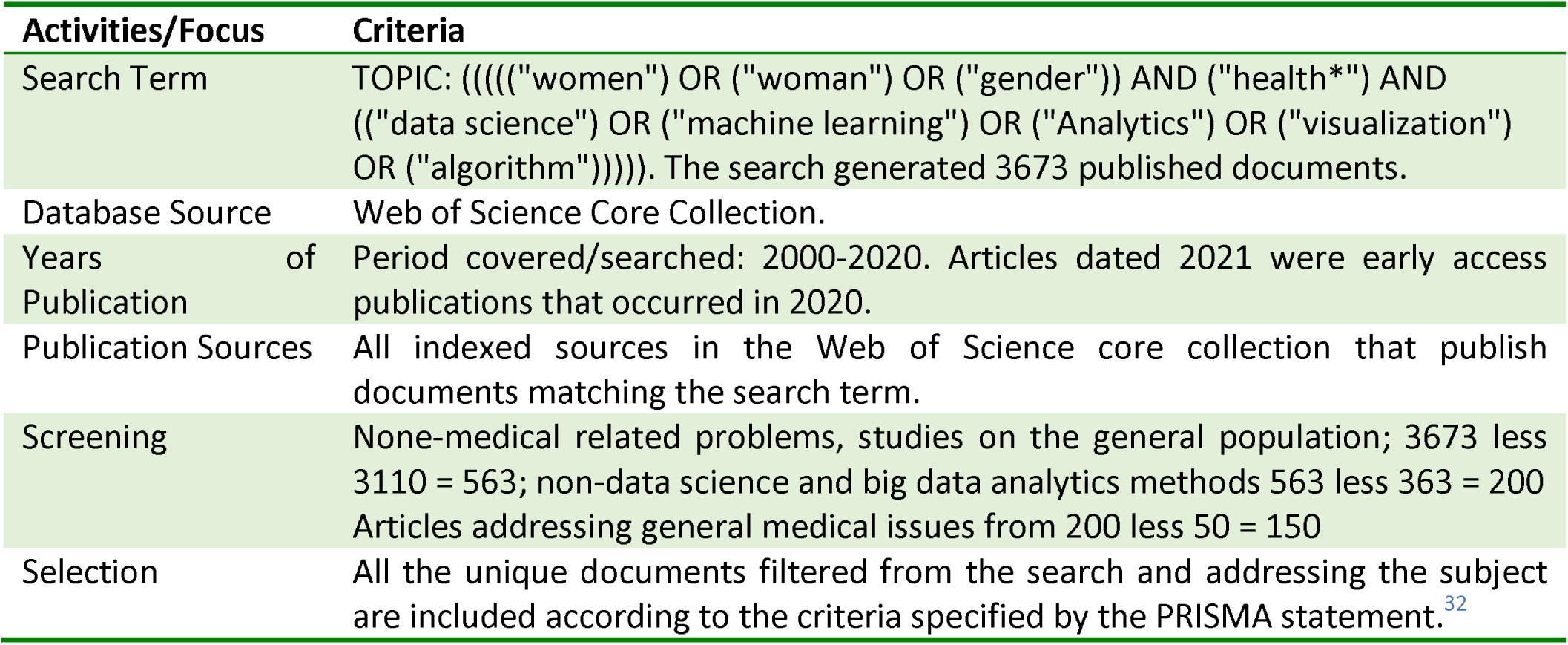
Articles’ Search, Filtering, Screening and Selection Criteria of Articles

### 3.3 Filtering, Screening and Selection of the Scientific Literature

The unique 3,673 published articles retrieved from the Web of Science/Knowledge went through a thorough filtering and screening process based on the criteria in Table 1. Finally, the study included 150 screened published documents. The next steps involved extracting the relevant information in textual form and exported to Excel and an R-studio programming environment for science mapping, quantitative analysis, and descriptive narratives.

### 3.4 Science Mapping and Visualization of Scientific Literature Using R-Base Open-Source Software

The analysis involved different mapping of DS techniques to diverse women’s health problems employs big data analytics and result visualization.^42,108^ The study also analyzed the authors, countries, and regions where the study originates and the researchers’ international collaborations. This helps to establish global awareness and utilization of modern DS techniques to improve women’s health worldwide. We utilize an open-source bibliometric analysis package (Bibliometrix) embedded in the R programming Studio to achieve these goals.^34^ The data used for the analysis are text files extracted from the 150 as explained in the above section. Further details about the open-source Bibliometrix and Vosviewer packages utilized for visualization are available elsewhere.^34,35^ The approach is suited to a new field or subject, such as contemporary DS and visual analytics.^11^

## 4. RESULTS

The summary of the data extracted from the published documents (Table 2) were generated using the R-programming Studio as described in the earlier section. The 150 papers included in the study appear in 113 different sources, indicating a broad range of publication outlets in diverse fields of research and practice. Also, 83.3% are published in journals, while 12% appear in the conference proceedings, both making 95.3% of the publications. The document per author (0.15) and the collaboration index (6.70) indicates a high collaboration among authors, which is expected in a multidisciplinary field (health informatics).

**Table 2.** Summary of the scientific literature production, authors, documents, and the citation impact

| Description | Results | Description | Results |
| --- | --- | --- | --- |
| Data coverage in years: 2000 to 2020; Active Period: 2006 to 2020 |  |  |  |
| Documents | 150 | Sources | 113 |
| Author Appearances | 1201 | Total Citation | 1078 |
| Authors/Single-authored documents | 4 | Average citations per documents | 7.19 |
| Authors – all authors/co-authors | 997 | Documents per Author | 0.15 |
| Authors of multi-authored documents | 993 | Co-Authors per Documents | 7.85 |
| Author Keywords | 493 | Authors Per Document | 6.52 |
| Collaboration Index | 6.7 | Keywords Plus (ID) | 544 |
| Types of Documents: |  |  |  |
| Articles (Journals) 83.3% | 125 | Papers in Conference Proceedings (12%) | 18 |
| Letters (1.3%) | 2 | Meeting Abstract (3.3%) | 5 |

### 4.1 Temporal trends of Data Science (DS) Methods Application in Women’s Health Research

The analysis in this Section intends to achieve the first research objective (RO1) as defined in the previous section.

#### 4.1.1 Annual Scientific Literature Production (SLP)

The SLP measures the development of a research field.36 The period 2000 to 2020 doubles as the emergence and growth of DS research and practice. The first article applying DS to solve Women’s health problems did not occur until 2006. The period 2006 to 2016 records just seven papers, while 143 SLP occurred from 2017-2020, indicating recent heightened interest (Figure 1). Studies using DS methods to solve women’s health make 26.6% during the period, while 73.4% relate to general health issues or problems relating to men (Table 1; Figure 1). The low document per author (0.15) – see Table 2, indicates fewer studies using DS techniques to address women’s health problems. Figure 1 shows a complete trend of the SLP during the period.

**Figure 1:**
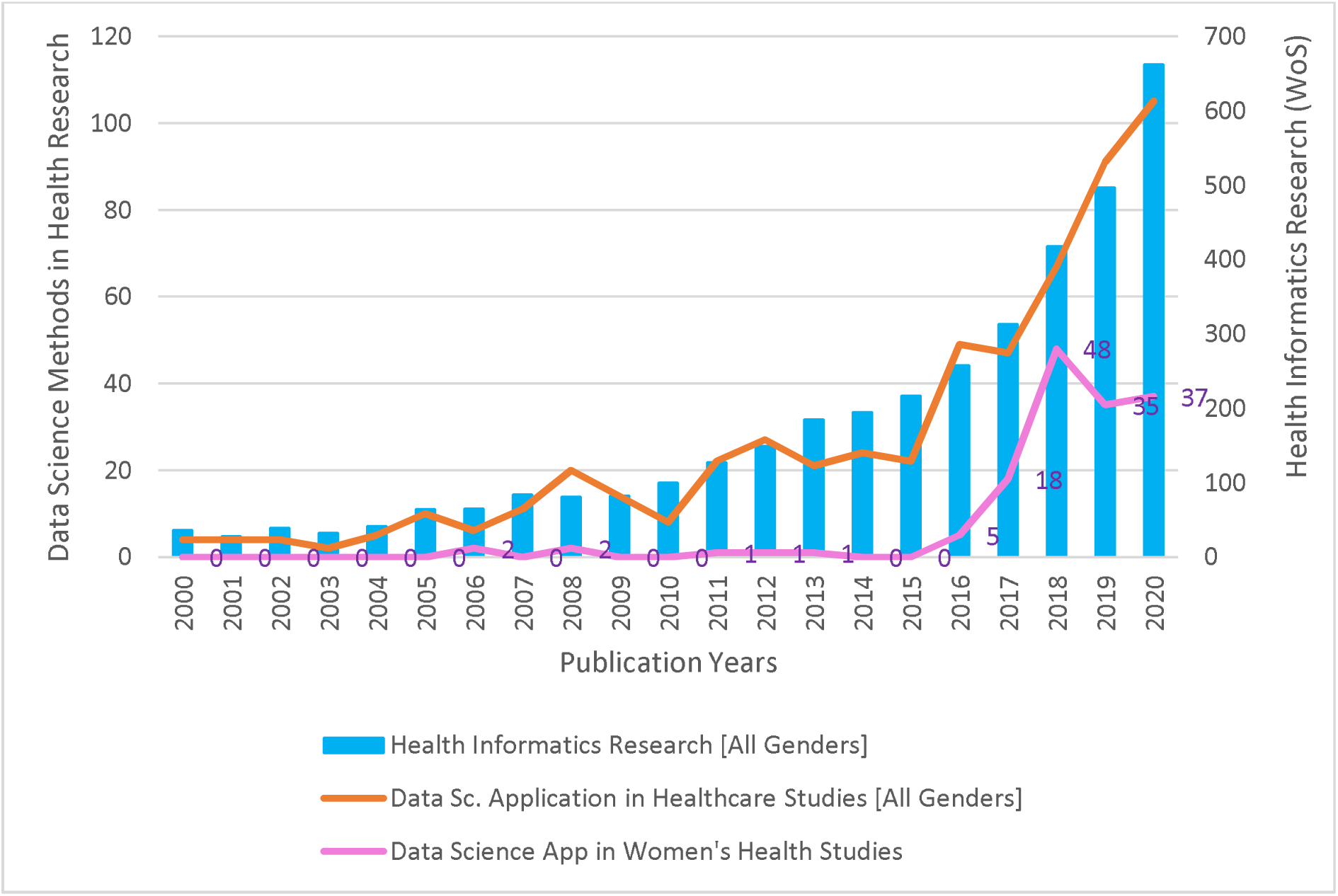
Annual scientific literature production on data science application in woman’s health

#### 4.1.2 Collaboration Network and Contributions of Countries and Institutions in DS in Women’s Health Studies

It is essential to know the institutions and countries pioneering the studies addressed in this paper as a pointer to the current spread and promoting more involvement. The 997 authors/co-authors come from 55 countries, with most of the authors domiciled in the USA (Table 2). Over 43% (65 out 150) of the SLP originate from research institutions in the USA, followed by India and Canada (Figure 2), both as corresponding authors within the USA (MCP) and co-authors with outside country collaborators (SCP)^11^. Figure 2 presents the top 20 countries.

**Figure 2.**
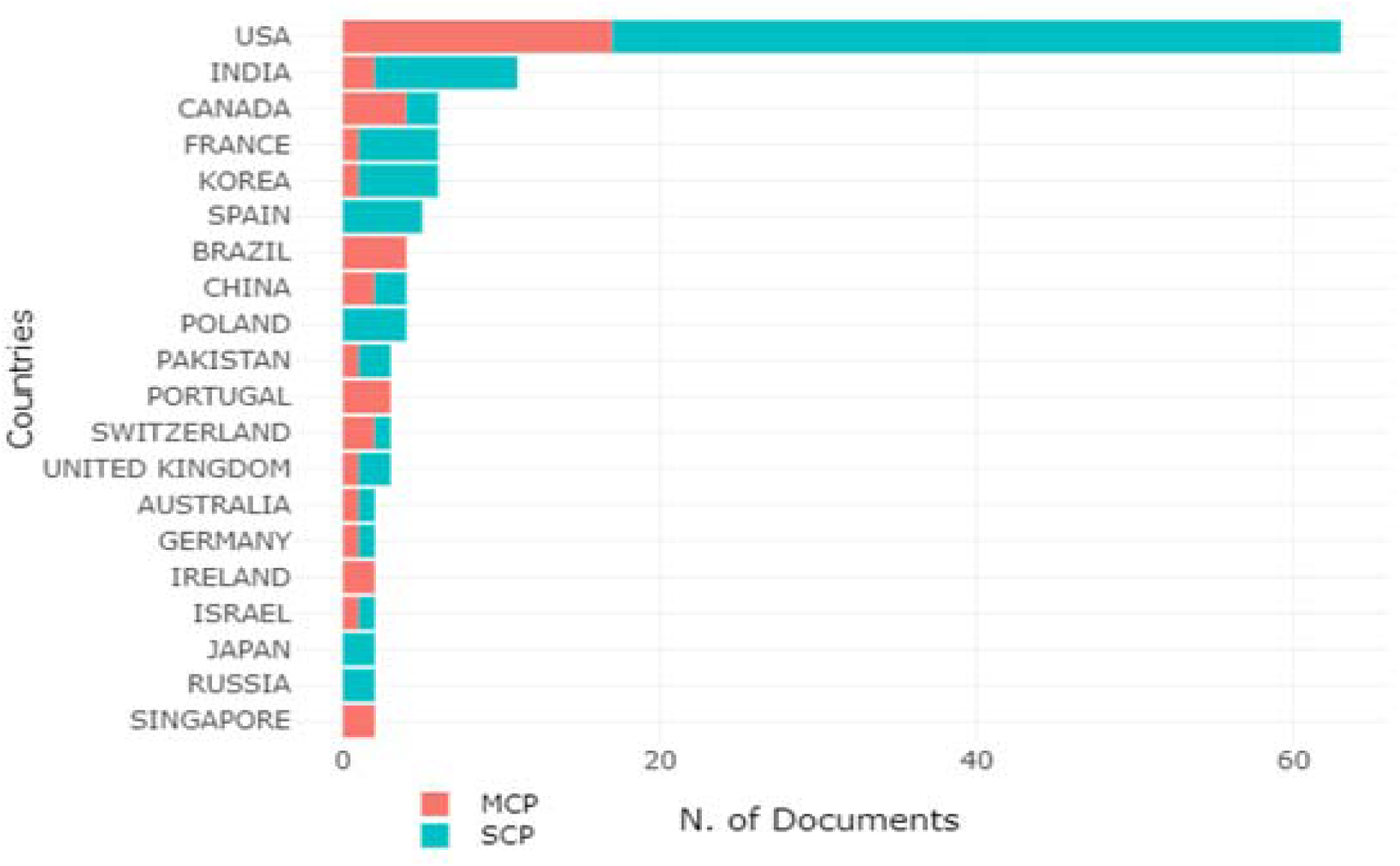
Countries Contributions of scientific literature and Collaborations (SCP: Corresponding Authors within the Country; MCP: Corresponding Authors Outside Country)

Regarding the contributing institutions, the most relevant and productive organizations originate from the USA (Figure 3), with the University of Washington, University of Pennsylvania, and Harvard Medical School, as the top three contributors. The nodes and the lines represent the contributions and connections, respectively.^59^ Figure 3 presents the complete lists of institutions from different countries.

**Figure 3.**
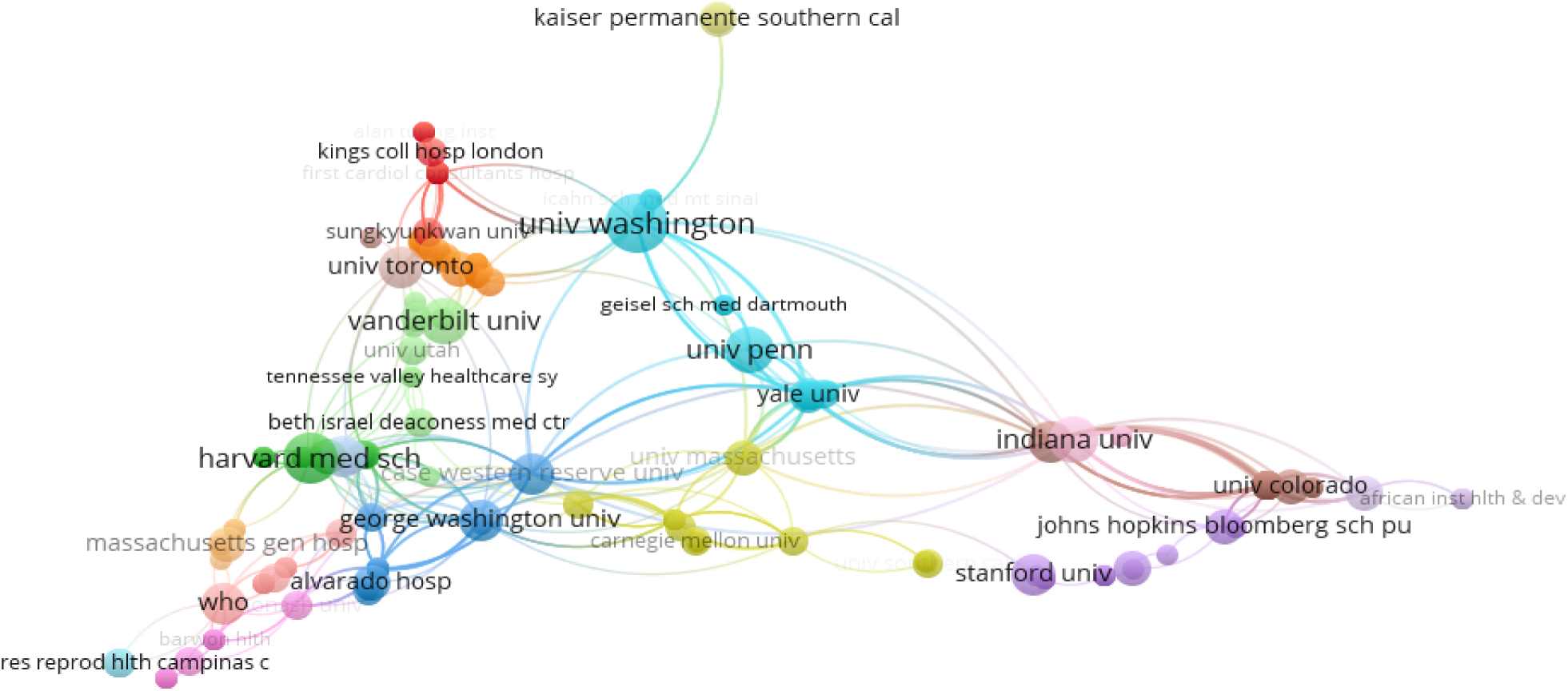
Institutions/Organizations involved in producing the scientific literature on data science application in women’s health conditions.

### 4.2 Science Mapping and Evaluation of Data Science Methods in Solving Women’s Health Problems

The analysis in this Section intends to achieve the first research objective (RO2).

#### 4.2.1 Science Mapping of Data Science Methods and Women’s Health Research, Authors, and Sources

Using an open-source bibliometric application embedded in R-Studio, we identify the top twenty-five (25) most frequent labels based on “authors’ keywords” as the unit of analysis.^33,59^ The authors’ keywords make up the central focus of any scientific literature (SL), as demonstrated by studies elsewhere.^31,45,59^ The result (Figure 4) shows machine learning as the most popular DS method used, while breast cancer, pregnancy issues, and other conditions, fall under gynecologic oncology.

**Figure 4.**
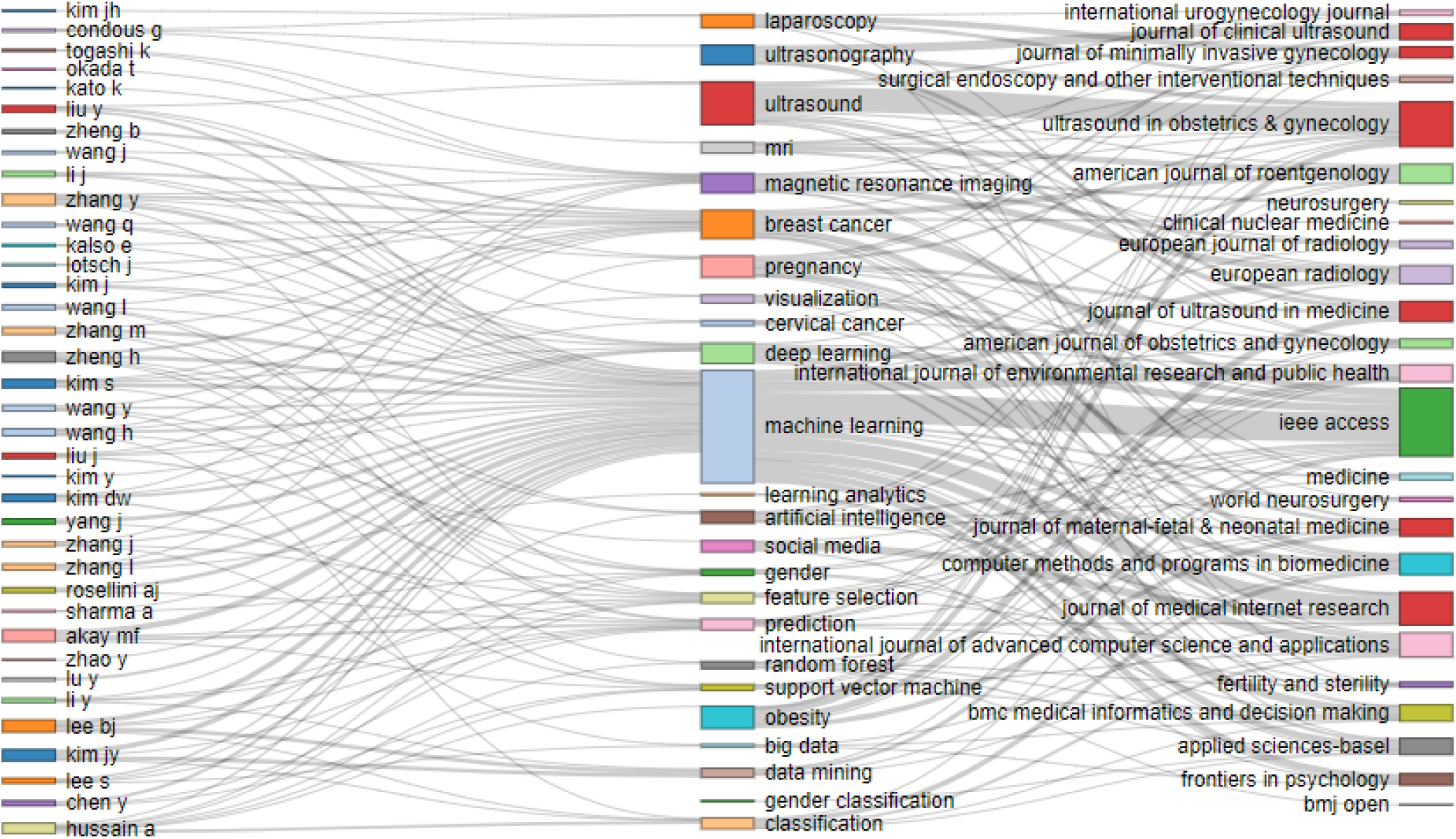
Top 25 data science methods and women’s health problems mapped to the authors and sources.

The efforts indicate a strategic approach in using DS to address health conditions that place the most health burden on women.^8,9^ On the sources, IEEE Access, Ultrasound in Obstetrics & Gynecology, JMIR publish most of the SLP on these subjects.

#### 4.2.2 Network Analysis and Visualization of DS Methods in Women’s Health Studies

The results of network analysis using VOSVIWER, an open-source science mapping application, demonstrate the connections between the DS techniques, the diseases examined, and the inter-relationships among infections and illnesses. For example, “machine learning” is commonly used to study breast cancer and pregnancy-related problems. Similarly, the ongoing SARS-CoV-2 condition indicates a connection with COVID-19 and cardiovascular diseases (Figure 5). The science mapping utilizes the 493 synthesized authors’ keywords (Table 2) as the unit of analysis and presents the interconnections and interrelationship among the terms stratified into clusters and produces the visualization.

**Figure 5.**
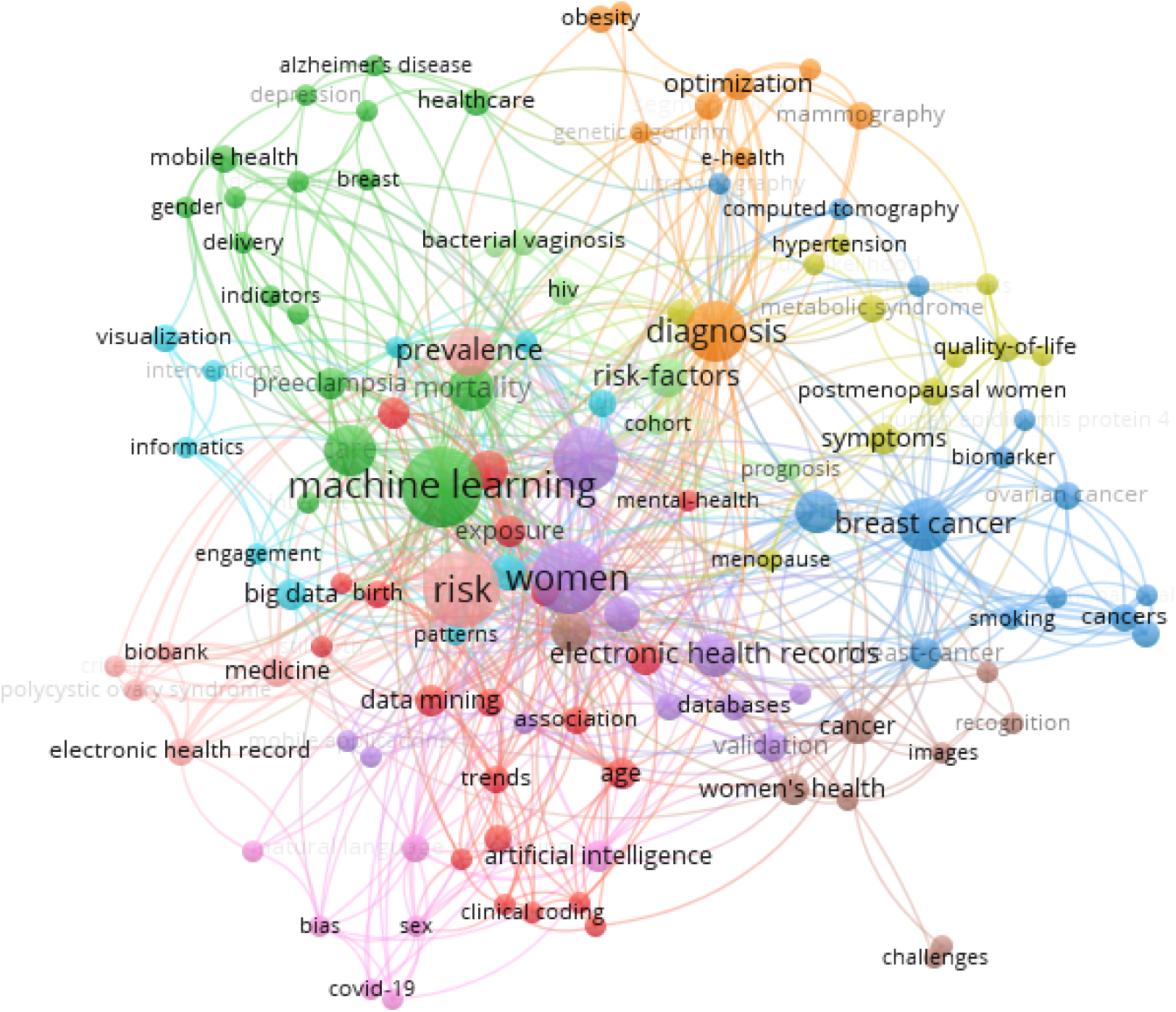
Visualization of the connections and inter-relationships among data science techniques and women’s health conditions.

#### 4.2.3 An Evaluation of Data Science Methods in Solving Women Health Problems

The 150 documents evaluated in this study employed eight (8) related data science (DS) methods including artificial intelligence (AI), machine learning (ML), deep learning (DL), and big data analytics (BDA). Others are predictive analytics (PA), novel algorithms (NA), clinical decision support systems (CDSS), and visual analytics (VA) (Table 3). The singular most frequently used data science method in solving women’s health problems is machine learning. Some studies use a combination of machine learning and novel algorithms, making it the most popularly used data science technique. Table 3 presents the complete results and descriptive analysis of the DS method and the problems tackled. These results echo the automatic mapping of the authors, keywords, and the sources (Figure 4/5) and network analysis that highlights the connections among the themes and topics using keywords as the unit if analysis (Figure 5).

**Table 3.**
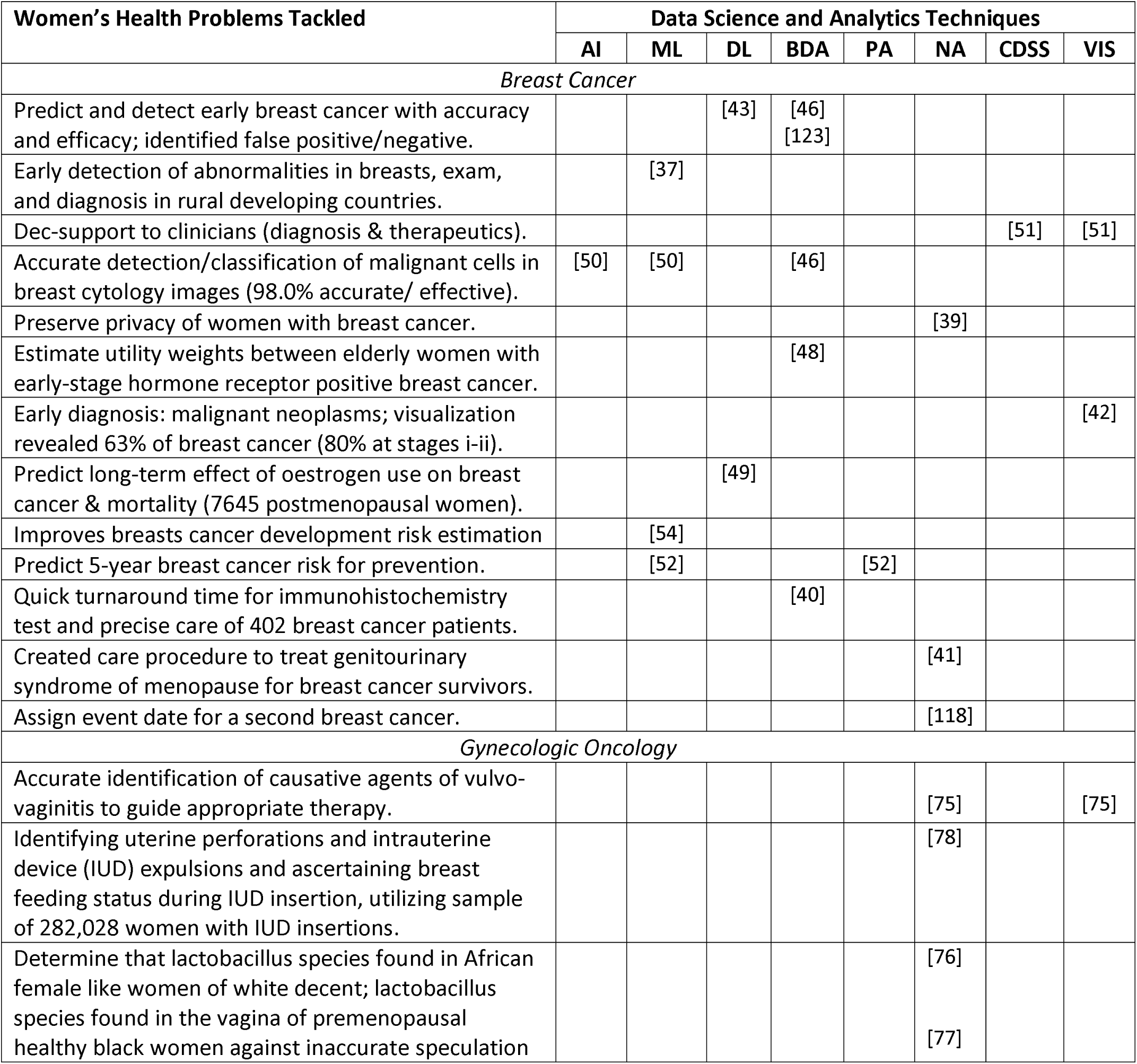
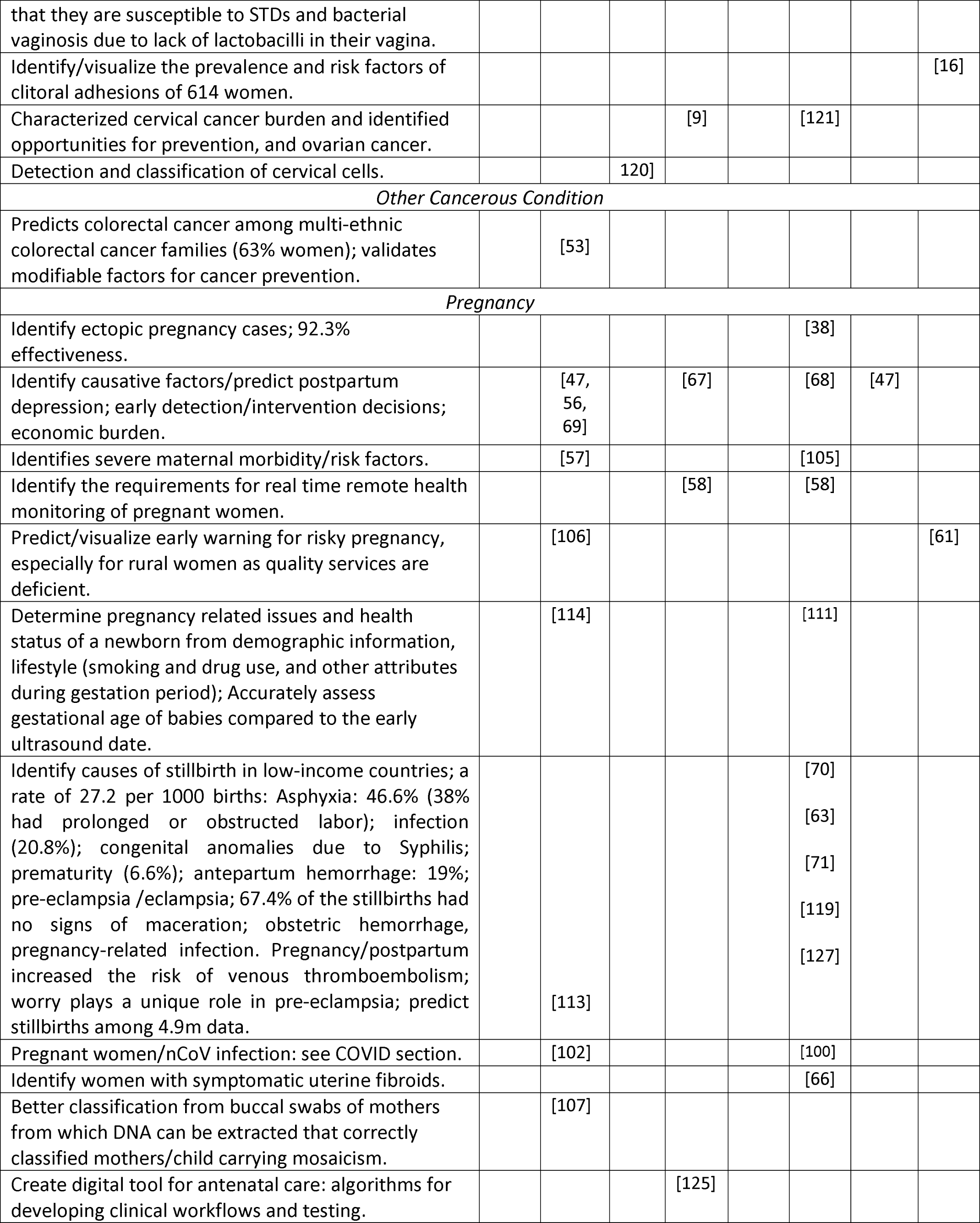
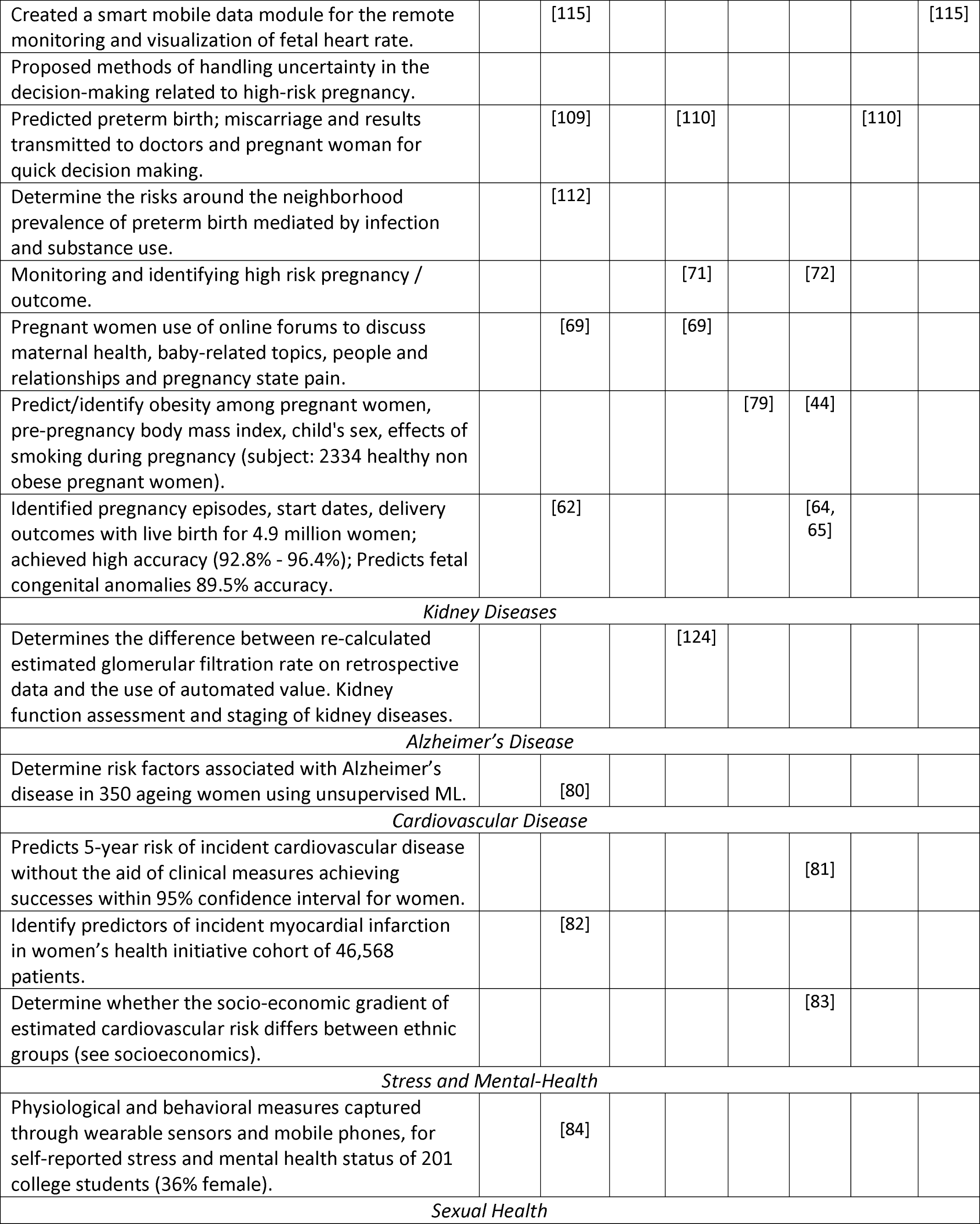
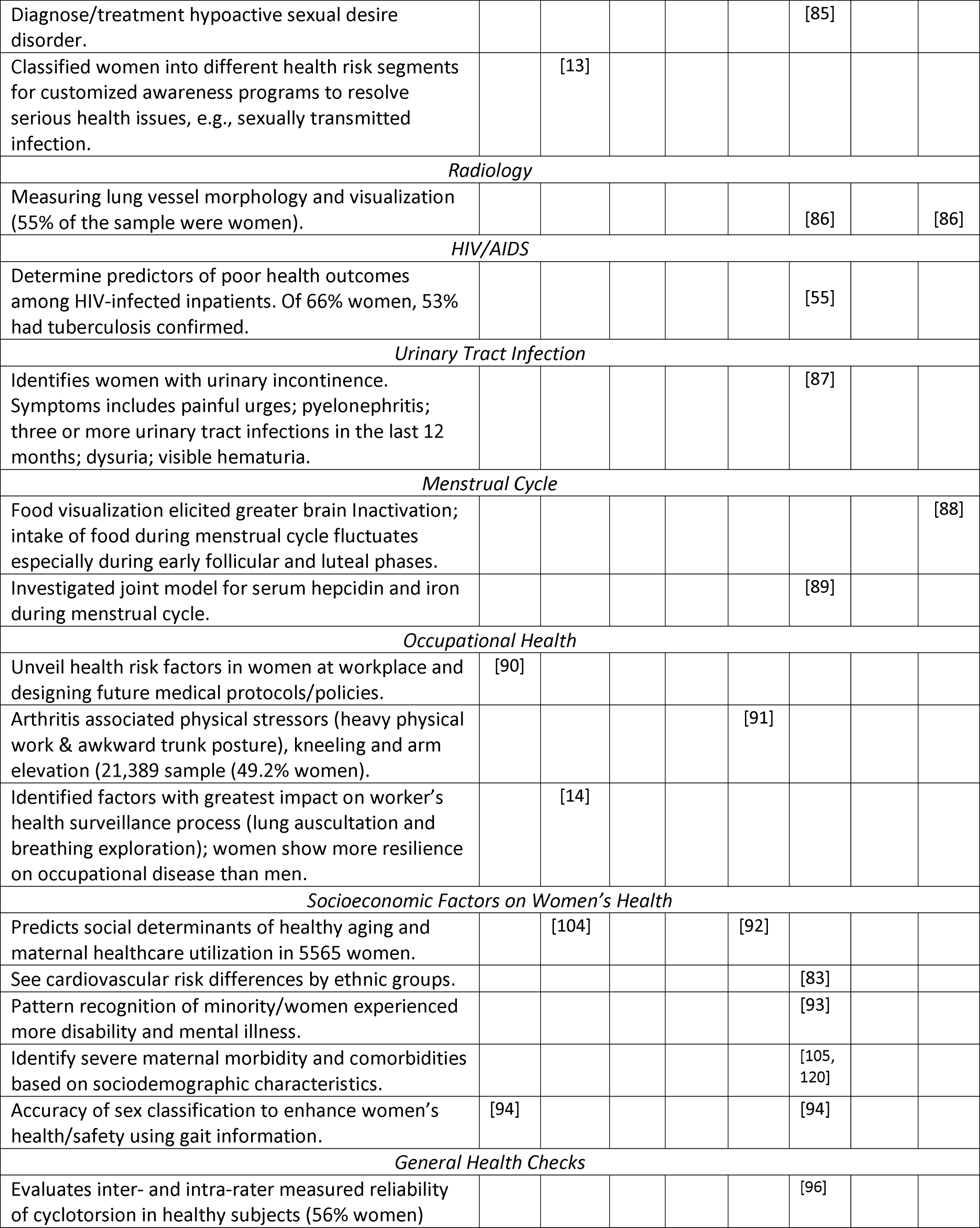
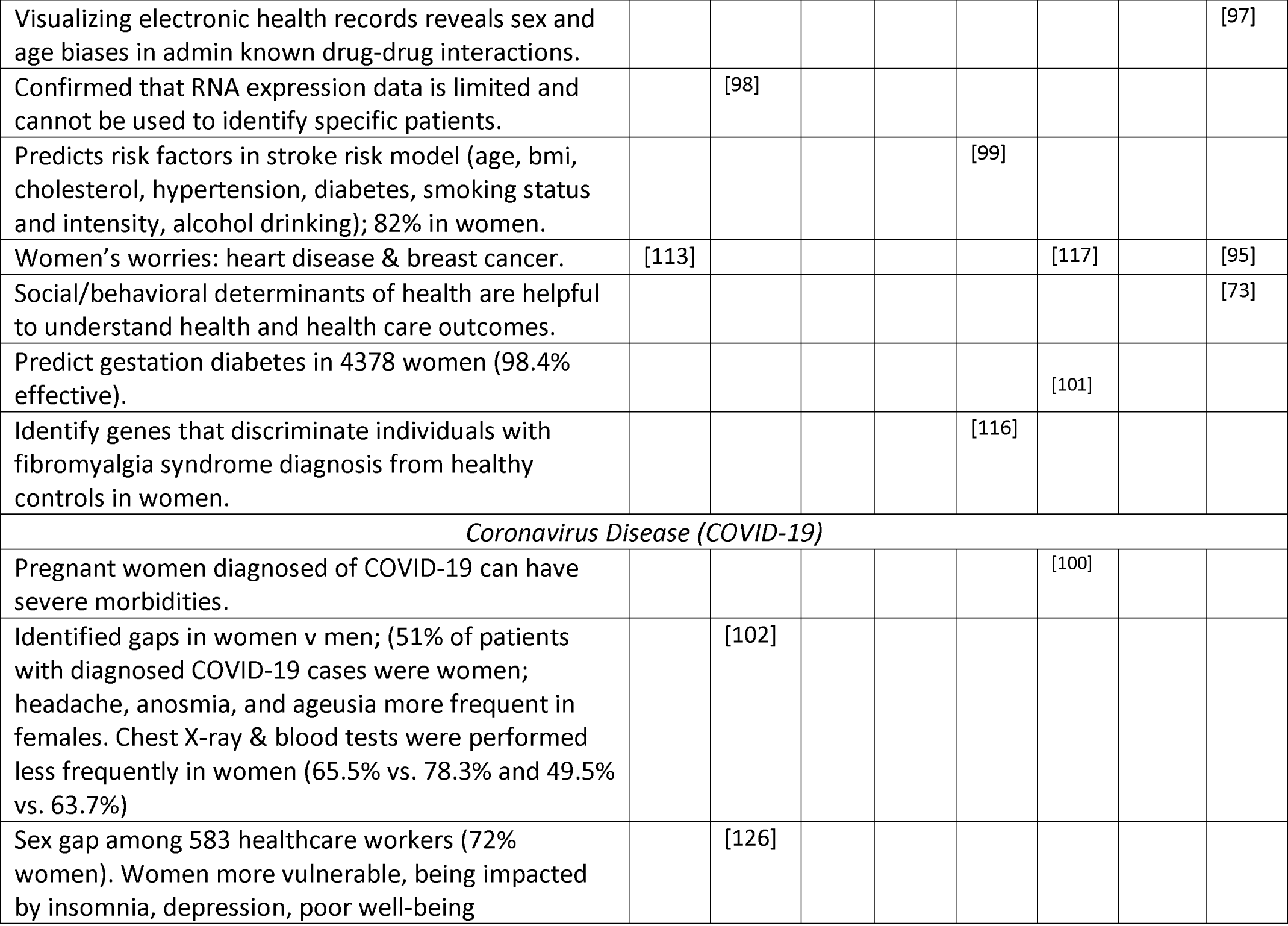
Data Science methods, problems tackled, and the application domain.

### 4.3 The Relationship Between the Scientific Literature (SL) and Usage and Citation Analysis

Here we examine the articles’ usage and citation impacts. The data from the selected 150 indexed documents on the WoS come in two streams, namely, U1 and U2 (Table 4).

**Table 4.** Usage and citation impact of the scientific literature on data science application in women’s health studies (U1, U2 = usage in the last 6 months, 7 years, respectively).

| REF | Article Focus | Sources | PY | Citation<br>(WoS) | U1 | U2 |
| --- | --- | --- | --- | --- | --- | --- |
| [49] | Breast cancer mortality | Lancet Oncology | 2012 | 221 | 0 | 25 |
| [76] | Lactobacillus vaginal microbiota | Sexually Transmitted Diseases | 2006 | 62 | 0 | 3 |
| [84] | Stress and mental health | J of medical Internet research | 2018 | 47 | 3 | 37 |
| [85] | Sexual desire disorder in women | Mayo Clinic Proceedings | 2018 | 36 | 2 | 14 |
| [88] | Brain responses to food images during menstrual cycle in healthy young women | American Journal of Clinical Nutrition | 2011 | 36 | 0 | 12 |
| [41] | Genitourinary syndrome of menopause in women with high risk for breast cancer | Menopause | 2018 | 36 | 1 | 8 |
| [100] | Care of pregnant women with COVID-19 in labor & delivery | American Journal of Obstetrics and Gynecology | 2020 | 27 | 6 | 8 |
| [119] | Syphilis during pregnancy and threat to maternal-fetal health | American journal of obstetrics and gynecology | 2017 | 25 | 0 | 17 |
| [104] | Factors determining healthy ageing | Scientific Reports | 2017 | 23 | 0 | 6 |
| [43] | Predicting breast cancer | Radiology | 2019 | 22 | 3 | 21 |
| [105] | Severe maternal morbidity and health disparities | Obstetrics and gynecology | 2017 | 22 | 0 | 7 |
| [70] | Stillbirth in low-and middle-income countries | International Journal of Obstetrics & Gynecology | 2018 | 22 | 0 | 5 |
| [79] | Organic pollutants and gestational diabetes among healthy US women. | Environment international | 2019 | 18 | 4 | 22 |
| [94] | Accurate sex classification to enhance women's health/safety. | Expert Systems with Applications | 2018 | 15 | 0 | 42 |
| [120] | Detection and classification of cervical cells | Journal of Supercomputing | 2020 | 15 | 18 | 18 |
| [81] | Predicts risk of cardiovascular disease for women. | CMAJ | 2018 | 13 | 0 | 3 |
| [50] | Detection and classification of breast cancer | Future Generation Computer Systems | 2019 | 11 | 0 | 5 |
| [63] | Maternal deaths in low / middle-income countries | International Journal of Obstetrics & Gynaecology | 2018 | 11 | 1 | 5 |
| [123] | Overcoming false positives and false diagnoses of breasts tumor | Journal of Grid Computing | 2018 | 10 | 0 | 18 |
| [101] | Gestational diabetes mellitus prediction in early pregnancy. | Scientific reports | 2019 | 10 | 0 | 7 |

- U1 (usage count within the past 180 days or last six months): 53 articles used 145 times.
- U2 (usage count in the past 7 years, 2014-2020): 121 articles used 806 times.

The results (U1=35.3%, U1=81%), indicating active usage of the scientific literature during the period shown. The data does not distinguish specific users (researchers, health/industry practitioners, and others) or for what purposes. The frequency of the articles’ usage also increased astronomically in the most recent years (U1=98%, U1=91% used between 2017-2020), indicating an active research area.

Regarding the citation impact, the total basic citation count was 1078, an average of 7.18 per document. However, 49 papers (32.7%) are yet to receive any citations, while 68.3% gained at least one citation, ranging as follows (1-10: 53.3%; 11-65: 14%; >65: <1% citations). Table 4 presents the top 20 most cited documents and the focus of women’s health research.

## 5. DISCUSSION

This study evaluates the DS methods employed in solving diverse women’s problems, including machine learning, big data analytics, deep learning, and novel/existing algorithms. Other techniques used are clinical decision support systems and visual analytics/visualization. These techniques were very effective in tackling diverse women’s health problems. Significant and improved health outcomes occurred in predicting and managing breast cancer, gynecologic oncology, and pregnancy-related health challenges (pre-eclampsia, eclampsia, prediction of risks during pregnancy, and more). Other health challenges addressed include menstrual cycle, psychosocial stress, gestational diabetes, and vaginosis. Diseases that are not solely related to women, like Alzheimer’s, cardiovascular diseases, and even scabies, are also examined. Occupational health and the importance of women’s health records confidentiality, and utilization of health facilities, family planning, and the characterization of drug-drug interaction. Table 3 presents a detailed analysis.

The research employing DS involves data collection through primary and secondary sources (medical records, claims data, and questionnaire administration) involving large volumes and sometimes complex data structures. The studies involved diverse groups of women, including patients, college students, healthcare and other workers, pregnant women, women living in a specified neighborhood, and patient caregivers. The sample sizes also vary for the different studies. The most extensive cohort study involved identifying pregnancy episodes and outcomes using a claims database of about 4.9 million women and the smallest that investigated brain activation during food visualization and the changes in estradiol concentration among nine women.^88,127^

A significant breakthrough in women’s health using DS occurred in using big data and visual analytics to undertake studies in obstetrics and gynecology, which helps to reduce maternal mortality.^11,128^ The methods use personal information such as sex and genomic data to predict cancer.^98^ However, there are ongoing challenges and societal hurdles, as some women seem uncomfortable using big data to develop health risk prediction.^129^ We believe that formulating appropriate privacy policies using big secondary data can resolve these issues.

The study also identifies disparities between women and minority groups. For example, a sex gap was identified in the treatment of COVID-19 among hospital patients. Although 51% of patients with diagnosed COVID-19 cases were women, more men were treated than women. Chest X-ray and blood tests were performed less frequently in women (65.5% vs. 78.3% and 49.5% vs. 63.7%).^102^ Issues relating to access to health are common, including women of different groups. Healthcare utilization differed across groups of cervical cancer patients, and this revelation can help identify opportunities for early prevention.^9^ A study found wealth, social group, literacy, religion, and early age at marriage as social determinants of maternal health care utilization.^92^ Midlife women’s health and analysis of most women’s concerns involves worries about breast cancer and heart disease.^95^

## 6. CONCLUSION

This study focused on evaluating DS methods and algorithms to solve women’s health problems. The goals in evaluating the techniques used, maps the methods and issues, analyzing the effectiveness, identifying what works, and what else can help to mitigate the disparities regarding women’s health burden and access to health. This paper has achieved these goals, further determines the origin and growth of this area of study, and predicts future growth using simple linear trends and forecasting.

The science mapping of the scientific literature production on this subject indicates an upward trend. About 95% (143 out of 150) of research utilizing data science methods in women’s health studies occurred within the last four (4) years (2017 to 2020), while only 4.7% of the research happened between 2000 and 2016, indicating a new field. The potency of the data science methods and the improved outcomes that can help solve women’s health problems raises hope of reducing the health gap between men and women, especially regarding the disease burden. However, addressing access to health requires more than advances in computer technology and the deployment of information systems and methods. Policymakers need to play a critical role in reducing the overall gaps, especially access to health. Further, the ratio of the studies involving data science to solving women’s health issues is relatively low and requires greater participation. Ironically, while the data science techniques appear promising to address women’s health problems, but some women activists are unknowingly opposing the adoption, indicating the need for enlightenment regarding the usefulness of the identified techniques.

## Data Availability

Not Applicable.

